# Physical activity, sedentary behaviour, and health inequalities among Somali residents in Sheffield, United Kingdom: a mixed-methods study

**DOI:** 10.64898/2026.05.18.26353489

**Authors:** Ayodele A. Falobi, Omar O Hersi, Opeolu O. Ojo

**Author notes:** **Corresponding Authors:** Dr. Opeolu Ojo, Director, Somaliland Institute for Non-Communicable Diseases, University of Burao, Burao, Somaliland and Head, Diabetes Research Group, University of Wolverhampton, United Kingdom, WV1 1LY.; **Additional Correspondence Author:** Dr. Ayodele Falobi, Somaliland Institute for Non-Communicable Diseases Training and Research (SINTRes), University of Burao, Burao, Somaliland.

## Abstract

**Background:** Physical inactivity and sedentary behaviour are major contributors to non-communicable diseases (NCDs) and are unevenly distributed across populations, disproportionately affecting migrants and ethnic minority groups. Somali communities in the UK experience multiple structural and socio-economic disadvantages; however, evidence on physical activity and associated inequities remains limited. This study examined physical activity, sedentary behaviour, and related barriers and facilitators among Somali residents in Sheffield, United Kingdom.

**Methods:** A cross-sectional mixed-methods study was conducted among Somali adults (n = 238). Quantitative data were collected using the International Physical Activity Questionnaire – Short Form (IPAQ-SF) and analysed using descriptive statistics and ordinal logistic regression. Qualitative data were obtained from two focus group discussions (n = 14) and analysed using inductive thematic analysis to explore socio-cultural, environmental, and structural determinants of physical activity.

**Results:** No statistically significant predictors of physical activity were identified in the adjusted analysis; however, consistent trends indicated lower activity levels among older adults and those in employment. Qualitative findings revealed multiple, intersecting barriers rooted in structural inequities, including migration-related lifestyle changes, reduced incidental activity, sedentary occupations, limited health literacy, language barriers, financial constraints, and gendered responsibilities. Cultural norms and environmental conditions further shaped behaviour. Facilitators included community-based, culturally tailored interventions, peer support, gender-sensitive programmes, and adaptation of traditional practices.

**Conclusion:** Somali residents in Sheffield face overlapping structural and socio-cultural barriers to physical activity that are not fully captured by quantitative measures alone. Equity-oriented, culturally competent, and community-led interventions addressing both systemic and behavioural determinants are essential to improve access to physical activity and reduce health inequalities and NCD risk.

## INTRODUCTION

Physical inactivity and prolonged sedentary behaviour are major modifiable risk factors contributing to the global burden of non-communicable diseases (NCDs), including cardiovascular disease, obesity, and type 2 diabetes mellitus (T2DM) (Lee et al. 2012; WHO, 2020). Insufficient physical activity is associated with increased risk of metabolic dysfunction, insulin resistance, and premature mortality, while regular physical activity plays a protective role in preventing and managing T2DM (Colberg et al. 2016). In Europe, approximately one in three adults do not meet recommended physical activity levels, and nearly half report no participation in structured exercise (OECD/WHO 2023). Sedentary behaviour, independent of physical activity, further exacerbates cardio-metabolic risk (Owen et al 2010). Importantly, these risk factors are socially patterned, disproportionately affecting socioeconomically disadvantaged and marginalised populations.

Migrant communities in high-income countries, particularly those originating from low- and middle-income regions such as sub-Saharan Africa, experience a disproportionate burden of NCD risk factors (Delavari et al. 2013). Although many migrants initially exhibit relatively favourable health profiles—a phenomenon described as the “healthy migrant effect”—this advantage often diminishes over time due to structural, environmental, and behavioural changes following migration. Evidence indicates that post-migration transitions are associated with reduced physical activity, increased sedentary behaviour, and adoption of less healthy lifestyles, contributing to widening health inequalities (Paudel et al 2025).

These behavioural shifts are shaped by interrelated structural and sociocultural determinants. Migration is frequently associated with a transition from physically active daily routines, including active transport and manual labour, to more sedentary occupational and domestic environments (Delavari et al. 2013). Environmental factors, such as colder climates, reduced daylight hours, and urban infrastructure that may not support accessible or culturally appropriate forms of physical activity, further constrain engagement (Ferguson et al. 2023). Socioeconomic disadvantage—including financial constraints, precarious employment, and limited access to recreational facilities—also reduces opportunities for active living (Hargreaves et al. 2019). In addition, language barriers, limited health literacy, and experiences of discrimination restrict access to health-promoting information and services (Ochieng 2013). These factors collectively reflect structural inequities that shape physical activity behaviours among migrant populations.

Within this broader context, Somali communities represent a significant and distinct population group in the United Kingdom. Somali migration has increased in recent decades, with over 100,000 individuals of Somali origin estimated to reside in the UK, primarily in urban centres including London, Birmingham, Manchester, and Sheffield (Jayaweera and Choudhury 2020, Office of National Statistics 2021). Many Somali migrants have refugee backgrounds, and their health outcomes are shaped by complex pre- and post-migration experiences, including displacement, socioeconomic disadvantage, and systemic inequalities (Im, Saleh and Khetarpal 2024).

Emerging evidence indicates low levels of physical activity and high sedentary behaviour among Somali populations in Europe. Studies have identified multiple barriers, including limited awareness of structured exercise, competing family responsibilities, lack of culturally appropriate facilities, and environmental constraints (Madar et al 2024, Andersen et al 2025). Cultural perceptions also play an important role; physical activity is often conceptualised as part of daily living rather than a discrete health behaviour, which may reduce engagement in structured exercise following migration to more sedentary environments (Andersen et al 2025). Gender norms may further restrict participation, particularly among women, due to preferences for gender-segregated environments and concerns around cultural appropriateness (Castaneda-Gameros, Redwood and Thompson 2018). These intersecting factors contribute to persistent inequalities in physical activity.

In the UK, Somali communities are disproportionately affected by socioeconomic disadvantage and barriers to healthcare access, increasing their vulnerability to NCDs (Public Health England 2018). Evidence suggests that Black African populations, including Somalis, have a higher risk of developing T2DM compared to the White British population, even at lower levels of adiposity (Agyeman and van den Born 2019). Physical inactivity and sedentary behaviour are likely to contribute significantly to this elevated risk within the context of broader structural determinants.

Despite these concerns, there remains a lack of community-specific data on physical activity and sedentary behaviour among Somali populations in the UK, particularly at the local level. Sheffield is home to a well-established Somali community and provides an important context for examining these issues. Generating context-specific evidence is essential for informing interventions that address health inequalities.

This study forms part of a broader initiative to develop culturally tailored interventions aimed at reducing health inequalities and preventing NCDs among Somali communities. Using Somali adults in Sheffield as a case study, the present study aims to assess levels of physical activity and sedentary behaviour and to identify associated barriers and facilitators. Understanding these determinants is critical for designing equity-oriented, culturally responsive interventions that address both behavioural and structural drivers of health.

## METHODS

### Study Design and Setting

This study employed a cross-sectional mixed-methods design, combining a quantitative survey with qualitative focus group discussions to examine physical activity, sedentary behaviour, and associated barriers among Somali adults in Sheffield, United Kingdom. A cross-sectional approach was used to provide a snapshot of behavioural patterns and associated determinants within a marginalised population at a single point in time [19]. The qualitative component was incorporated to enhance understanding of the socio-cultural and structural factors shaping physical activity behaviours and health inequalities.

Sheffield is a large urban city in the north of England with a well-established Somali community, estimated to comprise approximately 8,000–10,000 individuals. The study was conducted in collaboration with ISRAAC, a community-based organisation providing social, educational, and health-related support to Somali residents.

### Participants

The study population comprised Somali adults (aged ≥18 years) residing in Sheffield. Eligibility criteria included self-identification as Somali, residence within Sheffield, and ability to provide informed consent. Individuals with severe physical or cognitive impairments preventing participation were excluded.

### Quantitative Component

A priori sample size estimation was conducted using standard methods for cross-sectional studies (Charan and Biswas 2013). Assuming a prevalence of insufficient physical activity of 50%, a 95% confidence level, and a 5% margin of error, the required sample size was 244 participants. Participants were recruited using community-based convenience sampling in partnership with ISRAAC through community networks, social media, and engagement with community leaders and events. A total of 238 participants completed the survey, which was considered adequate for robust analysis. Data were collected using the validated International Physical Activity Questionnaire – Short Form (IPAQ-SF), administered online via Google Forms (Craig et al. 2003). The IPAQ-SF assesses frequency and duration of walking, moderate, and vigorous activities, as well as time spent sitting over the previous seven days. Additional items captured sociodemographic characteristics (age, gender, education, and employment), self-reported health status, and perceived barriers and facilitators to physical activity.

### Qualitative Component

To complement quantitative findings, two focus group discussions were conducted with 14 Somali participants (7 per group), purposively selected to ensure variation in age and gender. A semi-structured guide explored perceptions of physical activity, sedentary behaviour, cultural and environmental influences, and potential intervention strategies. Focus groups were moderated by a trained researcher in a culturally sensitive and supportive environment. Discussions were audio-recorded with participant consent.

### Data Analysis

#### Quantitative data

Descriptive statistics were used to summarise participant characteristics. Continuous variables were reported as means and standard deviations, and categorical variables as frequencies and percentages. Physical activity was assessed using the IPAQ-SF and scored according to established protocols (Craig et al. 2003). Activity data were converted into metabolic equivalent task minutes per week (MET-min/week) using standard values: walking (3.3 METs), moderate activity (4.0 METs), and vigorous activity (8.0 METs). Total physical activity was calculated as the sum across all domains. Participants were categorised into three levels: inactive (<600 MET-min/week), moderately active (≥600–<1500 MET-min/week or meeting minimum activity thresholds), and highly active (≥1500 MET-min/week through vigorous activity or ≥3000 MET-min/week overall).

Associations between physical activity levels and socio-demographic variables were examined using chi-square tests. Bivariate ordinal logistic regression analyses were conducted, and variables with P < 0.10 were included in a multivariable model. A parsimonious multivariable ordinal logistic regression model was fitted to identify independent predictors of physical activity. Model fit was assessed using −2 log likelihood and pseudo-R^2^ statistics (Cox and Snell; Nagelkerke), and the proportional odds assumption was evaluated using the test of parallel lines. Adjusted odds ratios (AORs) with 95% confidence intervals (CIs) were reported.

#### Qualitative data

Qualitative data were analysed using inductive thematic analysis. Audio recordings were transcribed verbatim and checked for accuracy. Coding was conducted iteratively, with codes grouped into categories and overarching themes reflecting barriers and facilitators to physical activity. To enhance analytical rigour, themes were reviewed for internal consistency and distinctiveness, and representative quotations were selected. Multiple readings of the data ensured depth of interpretation.

#### Integration of methods

Quantitative and qualitative findings were integrated at the interpretation stage to provide a comprehensive understanding of physical activity behaviours and their underlying determinants within the study population.

### Ethical Considerations

Ethical approval was obtained from the Life Sciences Ethics Committee at the University of Wolverhampton, UK. The study adhered to the principles of the Declaration of Helsinki. Participants received information about the study and provided informed consent prior to participation. Participation was voluntary, and participants could withdraw at any time. Data were collected anonymously, and confidentiality was maintained throughout. All data were securely stored and accessed only by the research team.

## RESULTS

### Participants’ Profile

The study included 238 participants (Table 1), with a higher proportion of females (61.8%) than males (38.2%). Most participants were aged 36–45 years (36.6%), followed by 26–35 years (21.4%) and 46–60 years (17.6%), while smaller proportions were aged 18–25 (13.4%) and over 60 years (10.9%). Nearly half had primary education (45.4%), with fewer reporting tertiary (31.9%) or secondary education (22.7%). Employment status was mixed, with 40.3% unemployed/retired, 39.5% employed, and 20.2% students. The majority were single or divorced (57.6%), and most had lived in the UK for over five years (73.9%)

**Table 1:** Demographic Characteristics and Quantification of Physical Activities and Sedentary Behaviour of Participants.

| Characteristics | Frequency<br>N (%) | Physical Activities<br>(MET-min/week) | Sedentary Time<br>(hr/day) |
| --- | --- | --- | --- |
| **Gender** |  |  |  |
| Male | 91 (38.2) | 921.3 ± 244.7 | 7.8 ± 1.3 |
| Female | 147 (61.8) | 515.7 ± 92.7 | 9.8 ± 2.1 |
| **Age (Years)** |  |  |  |
| 18 – 25 | 32 (13.4) | 1655.7 ± 247.3 | 7.2 ± 2.0 |
| 26 – 35 | 51 (21.4) | 930.5 ± 252.0 | 8.3 ± 1.9 |
| 36 – 45 | 87 (36.6) | 656.3 ± 152.3 | 8.7 ± 1.6 |
| 46 – 60 | 42 (17.6) | 185.7 ± 41.0 | 9.4 ± 1.5 |
| >60 | 26 (10.9) | 164.8 ± 36.2 | 10.4 ± 2.2 |
| **Education** |  |  |  |
| Primary | 108 (45.4) | 618.9 ± 47.7 | 10.0 ± 2.8 |
| Secondary | 54 (22.7) | 702.3 ± 252.3 | 8.5 ± 1.9 |
| Tertiary | 76 (31.9) | 834.5 ± 155.0 | 7.9 ± 1.8 |
| **Employment** |  |  |  |
| Student | 48 (20.2) | 1307.7 ± 347.2 | 8.1 ± 1.4 |
| Employed | 94 (39.5) | 691.9 ± 238.1 | 8.5 ± 1.9 |
| Unemployed/Retired | 96 (40.3) | 156.0 ± 29.3 | 9.8 ± 1.6 |
| **Marital** |  |  |  |
| Single/Divorced | 137 (57.6) | 612.0 ± 204.4 | 8.2 ± 1.5 |
| Married | 101 (42.4) | 825.0 ± 259.1 | 9.4 ± 1.9 |
| **Year in UK** |  |  |  |
| 0 – 5 years | 62 (26.1) | 660.0 ± 234.7 | 6.5 ± 1.8 |
| > 5 years | 176 (73.9) | 777.0 ± 226.0 | 9.1 ± 1.8 |
| Total | 238 (100) | 718.5 ± 156.9 | 8.8 ± 2.1 |

### Physical Activity Levels and Sedentary Behaviour

The mean total physical activity level among participants was 718.5 ± 156.9 MET-min/week (Table 1), indicating that, on average, the population met the threshold for moderate physical activity but did not reach levels consistent with high activity. As illustrated in Figure 1, a considerable proportion of participants were either inactive or only moderately active, reflecting generally suboptimal physical activity levels within the cohort (see also Supplementary Table 1).

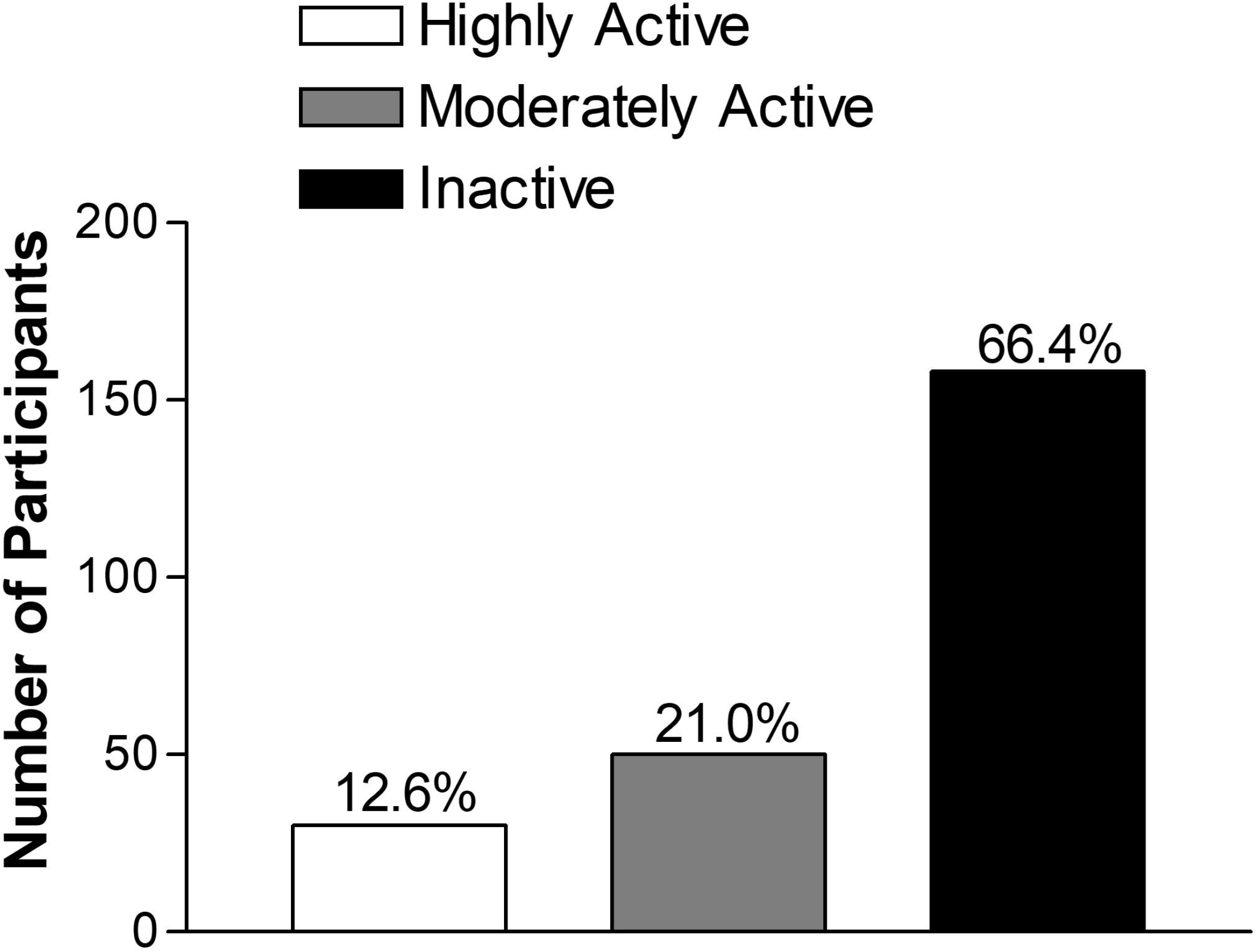

Marked differences were observed across demographic groups. Males reported substantially higher physical activity levels (921.3 ± 244.7 MET-min/week) compared to females (515.7 ± 92.7 MET-min/week). Physical activity declined with age, with the highest levels observed among participants aged 18–25 years (1655.7 ± 247.3 MET-min/week), followed by a progressive reduction across older age groups, reaching the lowest levels among those aged over 60 years (164.8 ± 36.2 MET-min/week)

Variations were also evident by education and employment status. Participants with tertiary education reported higher activity levels (834.5 ± 155.0 MET-min/week) compared to those with primary (618.9 ± 47.7 MET-min/week) or secondary education (702.3 ± 252.3 MET-min/week). Unemployed or retired individuals demonstrated markedly lower activity levels (156.0 ± 29.3 MET-min/week) compared to employed participants (691.9 ± 238.1 MET-min/week) and students (1307.7 ± 347.2 MET-min/week). Married participants exhibited higher activity levels than those who were single or divorced, and individuals who had resided in the UK for more than five years reported higher activity levels than more recent migrants

Sedentary behaviour was high across the sample, with a mean sitting time of 8.8 ± 2.1 hours/day. Females reported higher sedentary time (9.8 ± 2.1 hours/day) than males (7.8 ± 1.3 hours/day). Sedentary time increased with age, peaking among those aged over 60 years (10.4 ± 2.2 hours/day) (Table 1). Higher sedentary time was also observed among individuals with primary education (10.0 ± 2.8 hours/day) and among unemployed or retired participants (9.8 ± 1.6 hours/day), whereas students reported comparatively lower sitting time. Overall, these findings indicate substantial variation in both physical activity and sedentary behaviour across socio-demographic groups, highlighting population subgroups at greater risk of inactivity and prolonged sedentary exposure.

### Multivariable Analysis of Factors Associated with Physical Activity

In the multivariable ordinal logistic regression model, no socio-demographic variable was significantly associated with physical activity (PA) level at the 5% level. The proportional odds assumption was satisfied (p > 0.05), indicating that the model was appropriate. Age showed a consistent inverse relationship with PA. Compared with participants aged 18– 25 years, those aged 36–45 years (AOR = 0.21, 95% CI: 0.04–1.13, P = 0.069) and 46– 60 years (AOR = 0.15, 95% CI: 0.02–1.12, P = 0.064) had lower odds of being in a higher PA category, with a similar pattern observed among those aged over 60 years (Table 2).

**Table 2:** Multivariable Ordinal Logistic Regression Analysis of Factors Associated with Physical Activity Levels.

| **Variable** | **Adjusted OR** | **95% CI** | **P-value** |
| --- | --- | --- | --- |
| Gender (Female vs Male) | 2.36 | 0.67–8.29 | 0.182 |
| Age 26–35 vs 18–25 | 0.56 | 0.19–1.65 | 0.292 |
| Age 36–45 vs 18–25 | 0.21 | 0.04–1.13 | 0.069 |
| Age 46–60 vs 18–25 | 0.15 | 0.02–1.12 | 0.064 |
| Age >60 vs 18–25 | 0.20 | 0.03–1.57 | 0.125 |
| Employment (Employed vs Student) | 0.41 | 0.13–1.31 | 0.133 |
| Employment (Unemployed/Retired vs Student) | 0.47 | 0.09–2.32 | 0.352 |

Employment status demonstrated a comparable trend. Relative to students, employed participants (AOR = 0.41, 95% CI: 0.13–1.31, P = 0.133) and those unemployed or retired (AOR = 0.47, 95% CI: 0.09–2.32, P = 0.352) had reduced odds of higher PA levels. Females had higher odds of being in a higher PA category compared with males (AOR = 2.36, 95% CI: 0.67–8.29, P = 0.182), although this was not statistically significant.

### Qualitative Findings: Barriers and Facilitators to Physical Activity

Thematic analysis identified multiple barriers and facilitators influencing physical activity and diabetes prevention (Table 3). A dominant theme was the transition in lifestyle following migration, characterised by reduced physical activity and increased reliance on motorised transport. Participants described walking as a routine aspect of life in Somalia, whereas in the UK, driving was both a necessity and a symbol of social status, contributing to sedentary behaviour. Occupational patterns, particularly taxi driving, further reinforced inactivity.

**Table 3:**
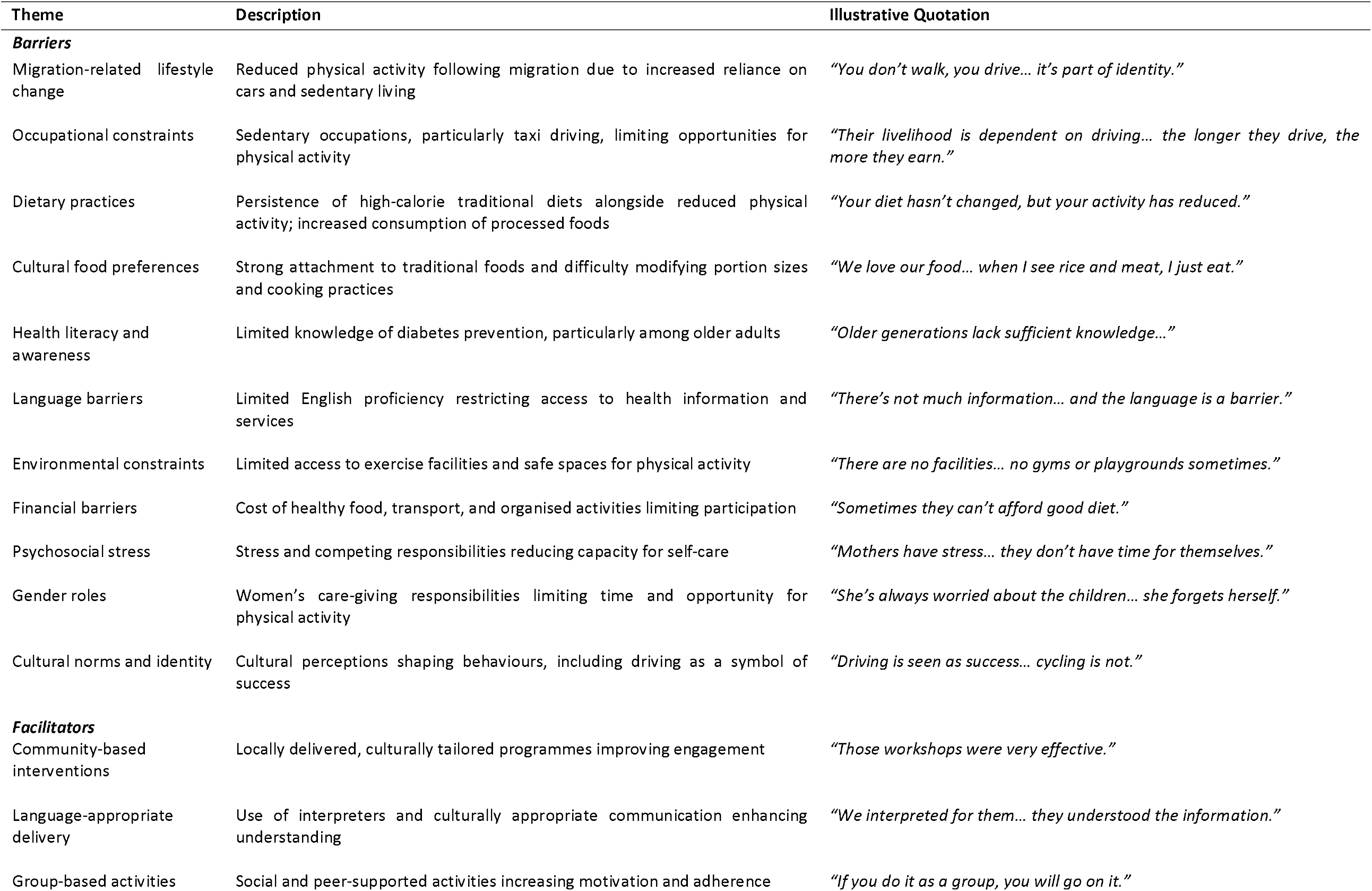
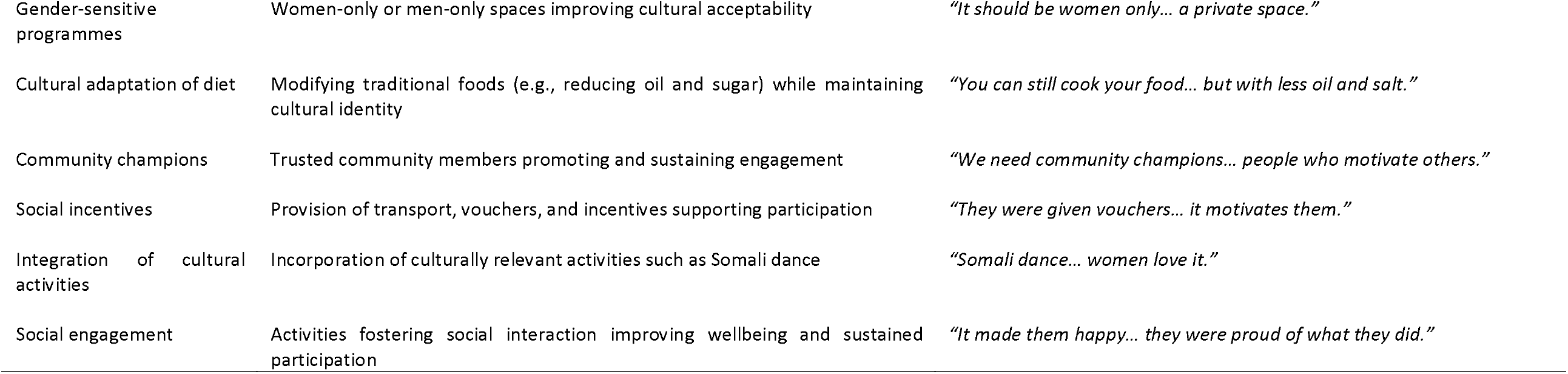
Summary of identified barriers and facilitators to physical activity among Somali adults in Sheffield.

| Theme | Description | Illustrative Quotation |
| --- | --- | --- |
| **Barriers** |  |  |
| Migration-related lifestyle change | Reduced physical activity following migration due to increased reliance on cars and sedentary living | *"You don't walk, you drive... it's part of identity."* |
| Occupational constraints | Sedentary occupations, particularly taxi driving, limiting opportunities for physical activity | *"Their livelihood is dependent on driving... the longer they drive, the more they earn."* |
| Dietary practices | Persistence of high-calorie traditional diets alongside reduced physical activity; increased consumption of processed foods | *"Your diet hasn't changed, but your activity has reduced."* |
| Cultural food preferences | Strong attachment to traditional foods and difficulty modifying portion sizes and cooking practices | *"We love our food... when I see rice and meat, I just eat."* |
| Health literacy and awareness | Limited knowledge of diabetes prevention, particularly among older adults | *"Older generations lack sufficient knowledge..."* |
| Language barriers | Limited English proficiency restricting access to health information and services | *"There's not much information... and the language is a barrier."* |
| Environmental constraints | Limited access to exercise facilities and safe spaces for physical activity | *"There are no facilities... no gyms or playgrounds sometimes."* |
| Financial barriers | Cost of healthy food, transport, and organised activities limiting participation | *"Sometimes they can't afford good diet."* |
| Psychosocial stress | Stress and competing responsibilities reducing capacity for self-care | *"Mothers have stress... they don't have time for themselves."* |
| Gender roles | Women's care-giving responsibilities limiting time and opportunity for physical activity | *"She's always worried about the children... she forgets herself."* |
| Cultural norms and identity | Cultural perceptions shaping behaviours, including driving as a symbol of success | *"Driving is seen as success... cycling is not."* |
| **Facilitators** |  |  |
| Community-based interventions | Locally delivered, culturally tailored programmes improving engagement | *"Those workshops were very effective."* |
| Language-appropriate delivery | Use of interpreters and culturally appropriate communication enhancing understanding | *"We interpreted for them... they understood the information."* |
| Group-based activities | Social and peer-supported activities increasing motivation and adherence | *"If you do it as a group, you will go on it."* |
| Gender-sensitive programmes | Women-only or men-only spaces improving cultural acceptability | *"It should be women only... a private space."* |
| Cultural adaptation of diet | Modifying traditional foods (e.g., reducing oil and sugar) while maintaining cultural identity | *"You can still cook your food... but with less oil and salt."* |
| Community champions | Trusted community members promoting and sustaining engagement | *"We need community champions... people who motivate others."* |
| Social incentives | Provision of transport, vouchers, and incentives supporting participation | *"They were given vouchers... it motivates them."* |
| Integration of cultural activities | Incorporation of culturally relevant activities such as Somali dance | *"Somali dance... women love it."* |
| Social engagement | Activities fostering social interaction improving wellbeing and sustained participation | *"It made them happy... they were proud of what they did."* |

Dietary practices emerged as a key barrier. Participants reported maintaining traditional high-calorie diets despite reduced physical activity, alongside increased consumption of processed and sugary foods. High sugar intake, particularly through tea, was described as culturally ingrained and difficult to modify. Limited health literacy and language barriers were also significant challenges, particularly among older adults. Participants highlighted a lack of culturally appropriate health information and limited engagement with healthcare services, which hindered awareness and prevention efforts. Structural and environmental constraints further limited healthy behaviours, including lack of accessible facilities, financial barriers, and competing demands such as childcare and employment. Short-term funding of interventions was identified as a barrier to sustainability. Psychosocial factors, particularly stress and gendered responsibilities, were prominent. Women described substantial care-giving burdens, limited time for self-care, and social isolation, all of which negatively impacted their ability to engage in physical activity.

Despite these barriers, several facilitators were identified. Participants emphasised the effectiveness of community-based, culturally tailored interventions, including group activities, peer support, and language-appropriate delivery. Women-only spaces and culturally relevant activities such as Somali dance were particularly valued. Practical strategies such as cooking demonstrations, promotion of healthier versions of traditional foods, and community-led initiatives were viewed as acceptable and effective. Social incentives, including transport support and small financial rewards, were also identified as important motivators.

## DISCUSSION

This study provides a comprehensive examination of physical activity (PA), sedentary behaviour, and associated determinants among Somali adults in Sheffield, integrating quantitative and qualitative findings to offer a nuanced understanding of behavioural patterns within a marginalised population. Although no statistically significant associations were identified in the multivariable analysis, consistent trends, supported by qualitative insights, highlight the role of structural, socio-cultural, and behavioural factors in shaping PA. Importantly, these findings underscore the social patterning of physical activity, reflecting broader health inequalities affecting migrant communities.

The quantitative findings indicated a non-significant trend of declining PA with increasing age, alongside lower activity levels among employed participants. These patterns are consistent with existing evidence demonstrating that PA declines with age and is influenced by occupational constraints and time limitations (Guthold et al. 2018). While the absence of statistical significance may reflect limited power, the direction of effects aligns with broader epidemiological evidence, suggesting that these trends are meaningful within the context of population-level inequalities.

Qualitative findings provided critical context, revealing that reduced PA is closely linked to migration-related lifestyle transitions. Participants described a shift from physically active routines in Somalia/Somaliland to more sedentary lifestyles in the UK, driven by changes in transport, occupational structures, and daily routines. This is consistent with evidence that migrants often experience declines in PA following resettlement due to environmental and socioeconomic changes (Delisle 2010, Osei-Kwasi et al. 2016). These transitions reflect structural constraints rather than individual choice, highlighting the role of broader social determinants in shaping behaviour. Such changes have important implications for metabolic health, as physical inactivity contributes to insulin resistance and increased risk of type 2 diabetes (WHO 2020).

Dietary practices further illustrate the interaction between behavioural and structural factors. Participants reported maintaining traditional high-calorie diets while experiencing reduced physical activity, alongside increased exposure to processed foods. This “nutrition transition” is well documented among migrant populations and is associated with increased risk of obesity and diabetes (Popkin 2017, Afshin et al. 2017). Importantly, participants emphasised the cultural significance of food and the difficulty of modifying long-standing practices, highlighting the need for culturally sensitive interventions that support adaptation rather than replacement.

Limited health literacy emerged as a key barrier to engagement in physical activity. Language barriers and lack of culturally appropriate information reduced awareness of PA benefits and restricted engagement with available services. These findings are consistent with evidence demonstrating that migrants with limited language proficiency have reduced access to health information and lower engagement in health-promoting behaviours (Berkman et al. 2017). These barriers reflect systemic inequities in access to health information, rather than individual deficits, underscoring the need for linguistically and culturally appropriate health promotion strategies.

Structural barriers—including financial constraints, limited access to facilities, and competing demands such as work and childcare—were also prominent. These findings align with socio-ecological models, which emphasise the influence of environmental and structural factors on health behaviours (Sallis, Owen and Fisher 2015). Participants also highlighted the short-term nature of many community programmes, suggesting that limited sustainability undermines long-term engagement. This reflects broader challenges in public health intervention design, particularly in underserved communities (Scheirer and Dearing 2011).

Psychosocial stress, particularly among women, emerged as an important determinant of PA. Participants described care-giving responsibilities, social isolation, and limited time for self-care, all of which constrained engagement in physical activity. These findings are consistent with evidence linking chronic stress to reduced PA and increased metabolic risk (Hackett and Steptoe 2017). The gendered nature of these experiences highlights the need for intersectional approaches that consider the combined effects of gender, migration, and socioeconomic disadvantage.

Cultural perceptions also played a dual role. While certain norms—such as viewing driving as a marker of success—acted as barriers, cultural identity also provided opportunities for engagement. Participants identified culturally relevant activities, such as Somali dance, as potential facilitators. This dual role of culture emphasises the importance of culturally informed and community-led approaches to intervention design (Ferguson et al 2023).

The study also identified several facilitators that can inform intervention development. Community-based, culturally tailored programmes were consistently viewed as effective, particularly when delivered in familiar and trusted settings. Peer support, group-based activities, and gender-sensitive approaches were important enablers of participation. Participants emphasised the value of adapting traditional practices—for example, modifying cooking methods—rather than replacing them. These findings are consistent with evidence demonstrating that culturally tailored interventions are more effective in promoting behaviour change among ethnic minority populations (Hawthorne et al. 2008). Addressing structural barriers, such as transport and cost, was also identified as essential for improving access.

The integration of quantitative and qualitative findings strengthens the interpretation of this study. While quantitative analyses did not identify statistically significant predictors, qualitative data provided critical explanatory insights into the mechanisms underlying observed patterns. This highlights the value of mixed methods approaches in capturing the complexity of health behaviours, particularly in marginalised populations where structural determinants play a central role.

## CONCLUSION

This study provides important insights into physical activity (PA) and sedentary behaviour among Somali adults in Sheffield, highlighting the influence of behavioural, structural, and socio-cultural determinants within a marginalised population. Although no statistically significant predictors were identified, consistent trends, supported by qualitative findings, indicate that increasing age, employment constraints, and migration-related lifestyle transitions contribute to lower PA levels. These findings underscore the social patterning of physical activity and the need to move beyond individual-level explanations.

Qualitative findings identified key barriers, including limited health literacy, language barriers, gendered responsibilities, financial constraints, and restricted access to appropriate facilities. At the same time, facilitators such as community-based, culturally tailored interventions, peer support, and gender-sensitive programmes were identified as effective. These findings highlight the importance of equity-oriented, community-led approaches that address both structural and socio-cultural determinants, supported by sustained investment and improved access to linguistically appropriate health information.

Several limitations should be acknowledged. The cross-sectional design limits causal inference, and the sample size may have reduced statistical power. Self-reported measures are subject to recall and social desirability bias, and recruitment through a community organisation may limit generalisability.

In conclusion, Somali adults in Sheffield experience intersecting structural and socio-cultural barriers to physical activity that contributes to health inequalities. Addressing these requires culturally competent, sustainable interventions that target systemic as well as behavioural determinants. Promoting equitable access to physical activity opportunities is essential to reduce disparities and the burden of non-communicable diseases in this population.

## ACKNOWLEDGEMENTS

We thank Mr. Adam Yusuf, ISRAAC Chairman and Mr Ahmed Mohamoud, Member of ISRAAC Board of Trustees for facilitating data collection.

## AUTHOR CONTRIBUTIONS

**OOO:** Writing – review & editing, Writing – original draft, Supervision, Project administration, Methodology, Investigation, Funding acquisition, Conceptualization; OOH: Writing – review & editing, Writing – original draft, Methodology, Conceptualization; AAF: Writing – review & editing, Writing – original draft, Software, Methodology, Formal analysis, Data curation, Conceptualization.

## DATA STATEMENT

Data for this project are available via https://doi.org/10.17605/OSF.IO/TDYHW

## ETHICAL STATEMENT

Ethical approval for the study was obtained from the Life Sciences Ethics Committee at the University of Wolverhampton, UK (LSEC/2022-23/OO/077).

## DECLARATION OF GENERATIVE AI AND AI-ASSISTED TECHNOLOGIES IN THE WRITING PROCESS

During the preparation of this work, the authors used ChatGPT to assist with language editing and improving clarity of expression. The AI tools were used solely to refine wording and structure; they were not used to generate data, conduct analyses, or influence the interpretation of results. All content was critically reviewed, revised, and approved by the authors, who take full responsibility for the integrity and accuracy of the manuscript.

## CONSENT FOR PUBLICATION

Not Applicable

## Tables

**Supplementary Table:**
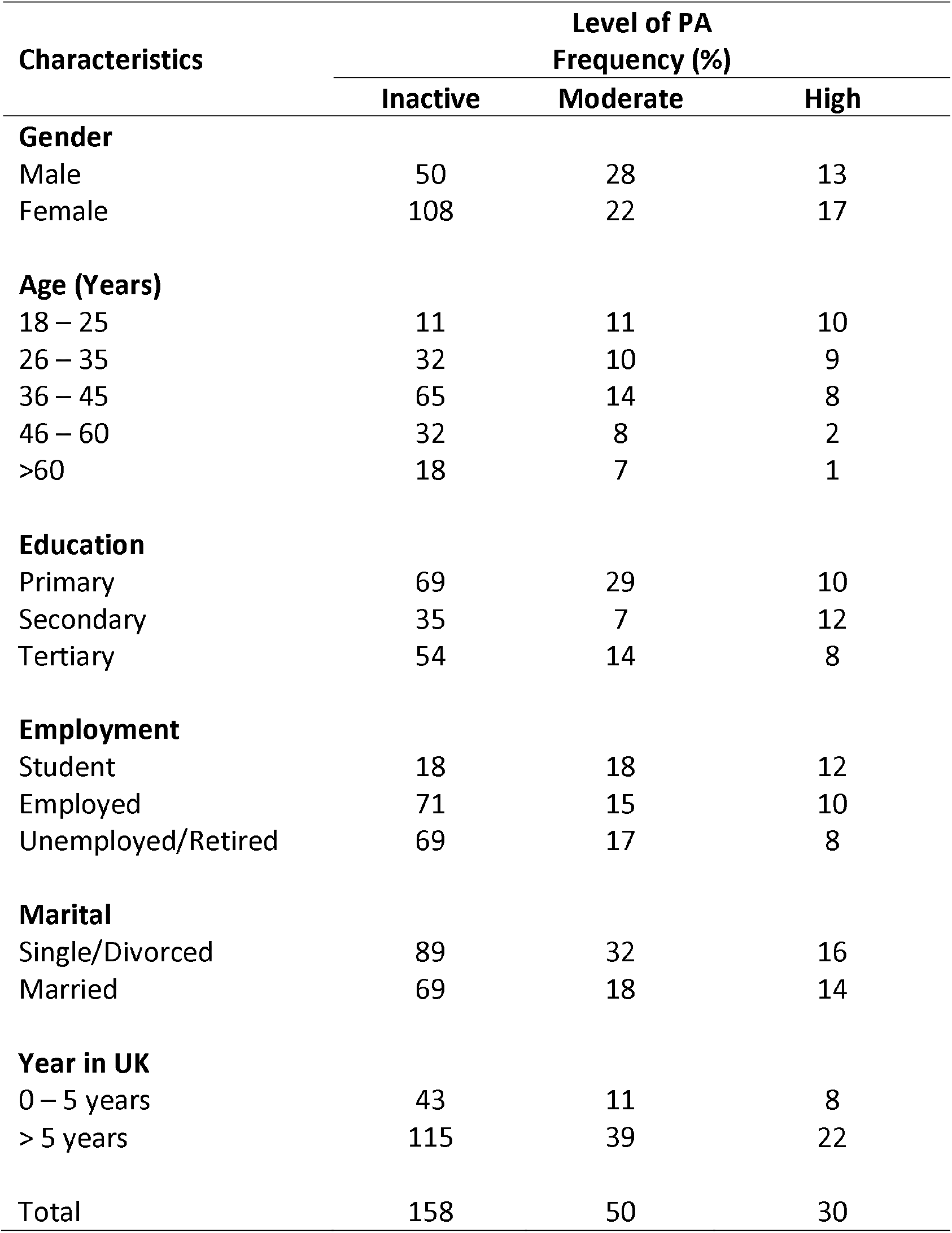
Level of PA in Participants.

| Characteristics | Level of PA<br>Frequency (%) |  |  |
| --- | --- | --- | --- |
|  | Inactive | Moderate | High |
| **Gender** |  |  |  |
| Male | 50 | 28 | 13 |
| Female | 108 | 22 | 17 |
| **Age (Years)** |  |  |  |
| 18 – 25 | 11 | 11 | 10 |
| 26 – 35 | 32 | 10 | 9 |
| 36 – 45 | 65 | 14 | 8 |
| 46 – 60 | 32 | 8 | 2 |
| >60 | 18 | 7 | 1 |
| **Education** |  |  |  |
| Primary | 69 | 29 | 10 |
| Secondary | 35 | 7 | 12 |
| Tertiary | 54 | 14 | 8 |
| **Employment** |  |  |  |
| Student | 18 | 18 | 12 |
| Employed | 71 | 15 | 10 |
| Unemployed/Retired | 69 | 17 | 8 |
| **Marital** |  |  |  |
| Single/Divorced | 89 | 32 | 16 |
| Married | 69 | 18 | 14 |
| **Year in UK** |  |  |  |
| 0 – 5 years | 43 | 11 | 8 |
| > 5 years | 115 | 39 | 22 |
| Total | 158 | 50 | 30 |

